# Programmatic nutritional support and tuberculosis treatment outcomes: a natural experiment in West Africa

**DOI:** 10.64898/2026.08.19.26360811

**Authors:** Mohammed Fall Dogo, Attannon Arnauld Fiogbe, Amanda Eng, Madolyn Dauphinais, Chelsie Cintron, Safiou Até, Christine Adjonou, Kokou Agossou, Meagan Karoly, Anne Fan Liu, Susie Jiaxing Pan, Marius Esse, Benjamin Adé, Komi Séraphin Adjoh, Dissou Affolabi, Akshay N. Gupte, Kobto G. Koura, Pranay Sinha

## Abstract

**BACKGROUND:** Undernutrition is the leading risk factor for tuberculosis (TB), yet evidence on programmatic nutritional support during treatment is limited. Benin and Togo are neighboring West African counties. Benin provides in-kind food support to all people with drug-susceptible TB; neighbouring Togo does not. This created the opportunity for a natural experiment.

**METHODS:** We conducted a prospective cohort study at 13 sites in Benin and Togo (September 2023-June 2024). We compared recipients of nutritional support with non-recipients, using Beninese non-recipients as an internal comparison. Primary outcomes were ≥5% weight gain at month 2, change in 6-minute walk test (6MWT) distance, and pill-count adherence. We used multivariable regression adjusted for pre-specified covariates.

**RESULTS:** Of 769 participants, 450 received nutritional support and 319 did not. Recipients had higher odds of ≥5% weight gain at month 2 (adjusted odds ratio [aOR] 1.57, 95% CI 1.13–2.19) and ≥10% at month 6 (aOR 1.92, 1.35–2.74), greater 6MWT improvement (adjusted β 40.6 m, 26.5–54.6), and higher adherence (aOR 3.43, 1.81– 6.51). Mortality was lower among recipients (aOR 0.32, 0.11–0.93). Sputum conversion and treatment success did not differ. Beninese non-recipients resembled Togolese participants across outcomes.

**CONCLUSION:** Programmatic nutritional support was associated with improved weight gain, functional recovery, adherence, and lower mortality during TB treatment, supporting its integration into national TB programmes.

## INTRODUCTION

Tuberculosis (TB) remains one of the leading infectious causes of mortality worldwide, with over 10 million incident cases and more than 1 million deaths annually [1].

Undernutrition impairs cell-mediated immunity against Mycobacterium tuberculosis. Undernourished persons with TB (PWTB) have increased disease severity, increased drug-related toxicity, and increased risk of unfavourable outcomes and mortality [2,3]. Despite this burden, nutritional support has received far less attention than HIV or diabetes as a modifiable risk factor in TB care [4]. A Cochrane review of macronutrient supplementation in PWTB found evidence supporting weight gain and accelerated sputum clearance but was limited by small sample sizes, heterogeneity, and insufficient power to detect effects on mortality [5]. Conducting well-powered randomised controlled trials of nutritional support is ethically challenging given the lack of equipoise, since it is difficult to justify withholding food from an already undernourished, vulnerable population [6]. As a result, the evidence base for programmatic nutritional support, particularly regarding mortality and treatment outcomes, remains thin and largely dependent on small randomised trials and non-randomised studies.

The INSTITUT (Impact of Nutritional Support for Tuberculosis on Intermediate and Terminal Undernutrition and Treatment Outcomes) study was designed to address this evidence gap by capitalising on a natural experiment in West Africa [7]. Benin’s national TB programme provides in-kind nutritional support to all PWTB as standard care, while Togo’s programme does not. This prospective cohort analysis compares outcomes across these two neighbouring countries, which share comparable demographics, socioeconomic conditions, TB incidence, and programme structures, to estimate the impact of programmatic nutritional support on nutritional recovery, functional capacity, and TB treatment outcomes.

## METHODS

### Study design and setting

We conducted a prospective cohort study at 13 treatment sites in Benin (6 sites) and Togo (7 sites) between September 2023 and June 2024. Benin and Togo are neighbouring West African countries with comparable population sizes, life expectancies, Gini indices (34.4 and 37.9) and TB incidence rates (52 and 32 per 100,000) [8]. Both have similarly structured national TB programmes with identical treatment protocols and receive support from the WHO and the Global Fund. Overlapping ethnic groups across the shared border contribute to comparable dietary practices. Benin’s national TB programme provides in-kind nutritional support to all people with drug-susceptible TB; Togo’s does not. This difference in programmatic policy defined a natural experiment. The study was registered (NCT06084715) and reported following STROBE.

### Participants

Adults aged 18 years or older with drug-susceptible, GeneXpert MTB/RIF-positive or smear-positive (≥1+ acid-fast bacilli) pulmonary TB receiving care at programme basic management units and willing to attend follow-up visits were eligible. We excluded people with drug-resistant TB, those who had received 7 or more days of antimicrobial therapy, those too unwell to enrol (WHO Performance Status 4), pregnant individuals, and those unable to complete the 6-minute walk test because of pre-existing mobility limitations.. Ethics committees in Benin and Togo approved the study; all participants provided written informed consent.Participants were followed until 6 months after treatment completion

### Exposure and intervention

The exposure was individual receipt of programmatic nutritional support, defined as any documented evidence of receiving the food basket or prepared meals (either quantified basket contents or a participant-reported record of receipt at the month 2 visit). Because Benin’s programme provides support to all patients while Togo’s provides none, recipients were drawn from Benin, and non-recipients comprised all Togo participants together with the Benin participants who did not receive support. Non-receipt among Beninese participants resulted from supply interruptions and stock-outs linked to Global Fund financing disruptions, not from clinical or individual selection. Benin’s programme, funded jointly by the Global Fund and the Government of Benin, provides a take-home food basket of spaghetti, corned beef, cassava flour, condensed milk, sardines, sugar, beans, and sorghum, given approximately monthly to outpatients during the intensive phase (Supplementary Figure 1). Hospitalised patients received three prepared meals daily. The type, quantity, and frequency of support varied by availability and site.

Beninese patients also received group nutritional counselling at follow-up visits.

### Variables and measurement

Pre-specified primary outcomes were weight gain of at least 5% at month 2, change in 6-minute walk test (6MWT) distance at month 6, and treatment adherence. Weight was measured at enrolment, month 2, and month 6 using standardised scales. The 6MWT was performed following the American Thoracic Society protocol [9]. Adherence was determined objectively by pill count at the end of the intensive phase and classified as adherent or non-adherent. Secondary outcomes were weight gain of at least 10% at month 6, sputum smear conversion at month 2 among those smear-positive at baseline, treatment success (cure or completion per WHO definitions), and all-cause mortality during treatment. Covariates included age, sex, multidimensional poverty [10], HIV status, diabetes, tobacco use, baseline body mass index, cavitation, and pleural effusion, ascertained at enrolment by interview, clinical examination, chest radiography, and laboratory testing.

### Bias

The principal concern was confounding by country-level differences between Benin and Togo. To address this, we compared three groups: Benin recipients, Benin non-recipients, and Togo participants. The Benin non-recipients, who shared Benin’s health system and sites but received no food support, served as an internal comparison to distinguish the effect of nutritional support from country-level factors. We report baseline characteristics of all three groups to allow assessment of residual confounding.

### Study size

The study was designed to detect a difference in a composite unfavourable outcome with 80% power at a 5% significance level. Enrolment did not reach the planned target during the study period, and the achieved sample size (769 participants) was below the design target of 1,100 participants described previously [7]. The study was therefore underpowered for the less frequent treatment outcomes.

### Statistical analysis

We compared recipients and non-recipients using logistic regression for binary outcomes and linear regression for change in 6MWT distance. Continuous variables were analysed on their original scale. Covariates were selected from univariable screening (p≤0.20) together with pre-specified clinical variables, and are listed for each model in the table footnotes. We report adjusted odds ratios (aOR) or regression coefficients (aβ) with 95% confidence intervals. Participants with missing outcome data were excluded from that analysis; missingness is reported by group (Supplementary Table 3). Analyses of sputum conversion, treatment success, and mortality involved few events and are reported as exploratory. No correction for multiple comparisons was applied. Because baseline 6-minute walk distance differed between groups and was inversely correlated with subsequent change, models for walk distance included baseline distance as a covariate; this specification is equivalent to analysis of covariance on the month 6 value and accounts for regression to the mean.

## RESULTS

### Participants

Of 1,188 patients assessed, 855 were eligible and 786 enrolled. Seventeen transferred out before outcomes could be ascertained, leaving 769 for analysis (Figure 1). Of these, 450 received nutritional support (all in Benin) and 319 did not (270 in Togo and 49 in Benin). Baseline characteristics are shown in Table 1. Non-recipients had more HIV (16.9% vs 7.4%), cavitary disease (81.9% vs 69.8%), and pleural effusion (15.2% vs 8.3%), whereas recipients had more hazardous alcohol use (41.1% vs 26.3%) and lower baseline BMI (18.9 vs 19.7 kg/m²). These imbalances were addressed through multivariable adjustment; the three groups differed on several characteristics (Supplementary Table 2).

**Figure 1.**
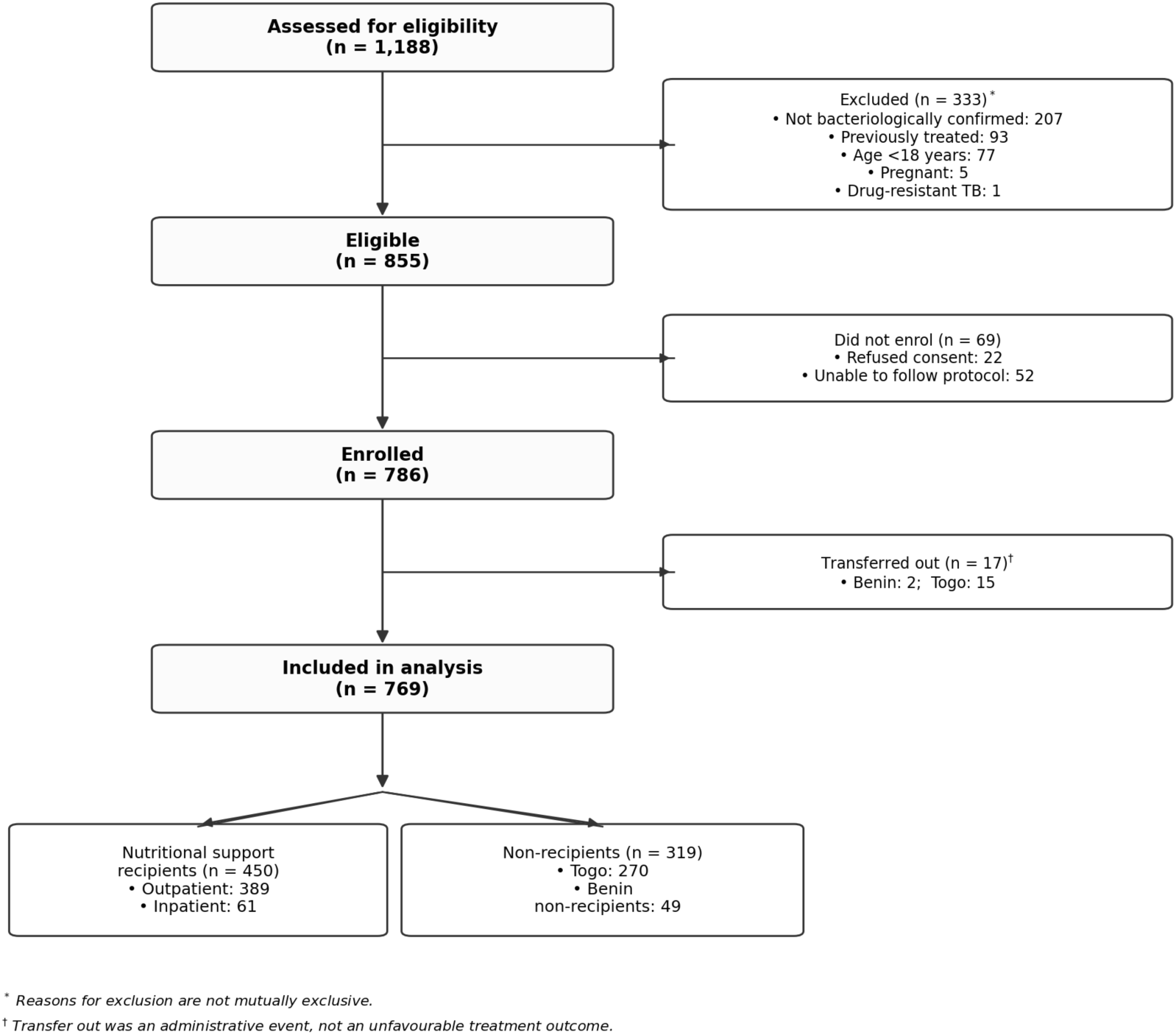
Participant flow. Of 1,188 patients assessed for eligibility, 769 were included in the analysis. Abbreviations: TB, tuberculosis.

**Table 1.**
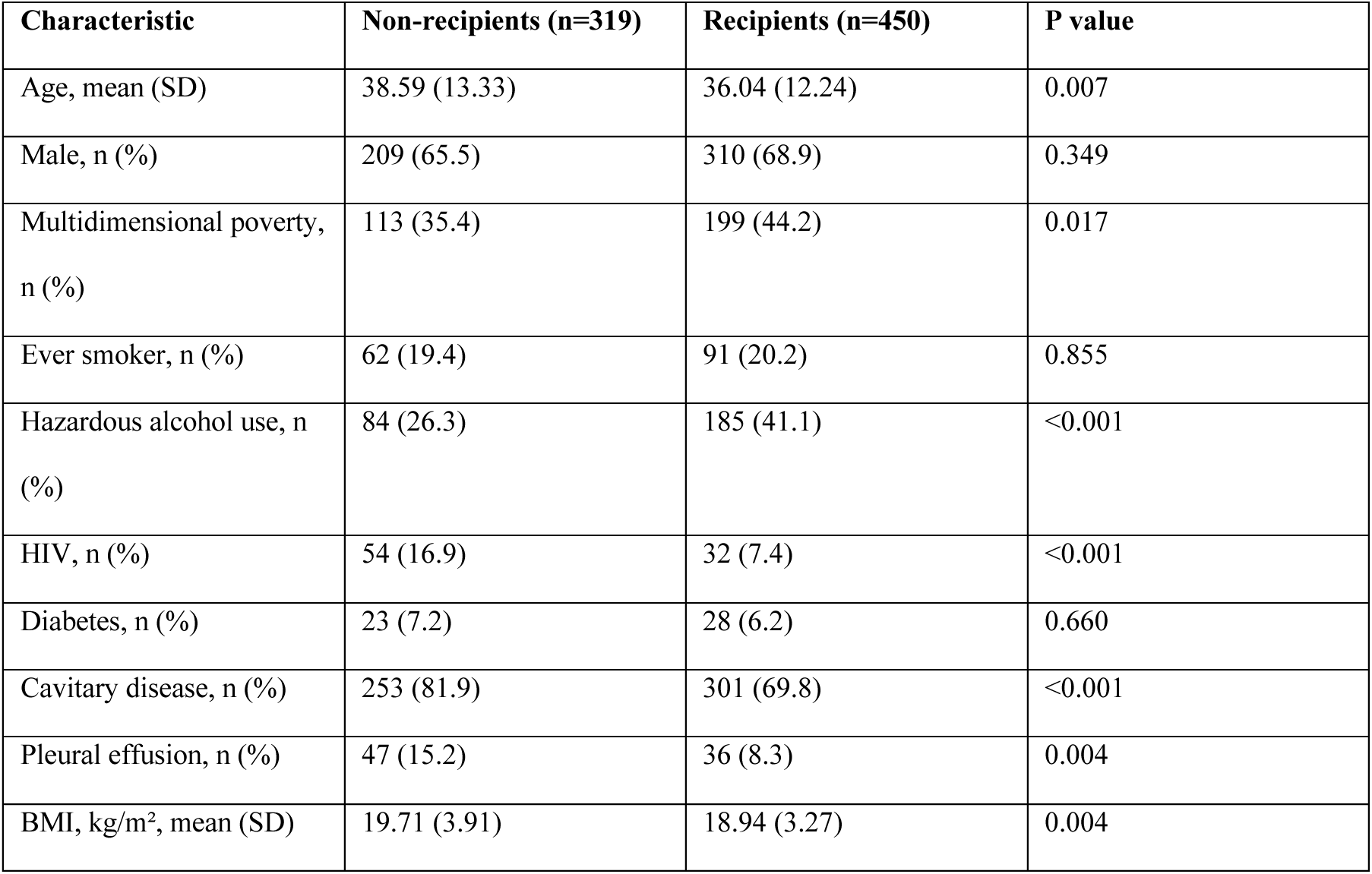
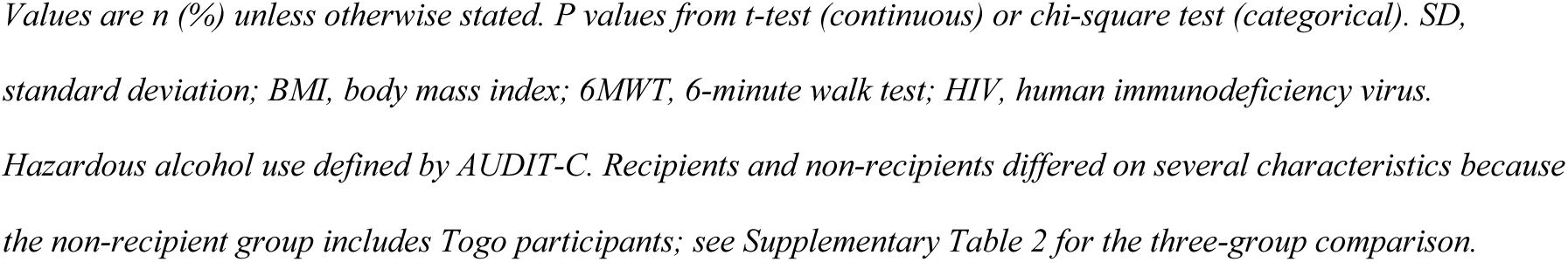
Baseline characteristics of participants by receipt of nutritional support.

| **Characteristic** | **Non-recipients (n=319)** | **Recipients (n=450)** | **P value** |
| --- | --- | --- | --- |
| Age, mean (SD) | 38.59 (13.33) | 36.04 (12.24) | 0.007 |
| Male, n (%) | 209 (65.5) | 310 (68.9) | 0.349 |
| Multidimensional poverty, n (%) | 113 (35.4) | 199 (44.2) | 0.017 |
| Ever smoker, n (%) | 62 (19.4) | 91 (20.2) | 0.855 |
| Hazardous alcohol use, n (%) | 84 (26.3) | 185 (41.1) | <0.001 |
| HIV, n (%) | 54 (16.9) | 32 (7.4) | <0.001 |
| Diabetes, n (%) | 23 (7.2) | 28 (6.2) | 0.660 |
| Cavitary disease, n (%) | 253 (81.9) | 301 (69.8) | <0.001 |
| Pleural effusion, n (%) | 47 (15.2) | 36 (8.3) | 0.004 |
| BMI, kg/m<sup>2</sup>, mean (SD) | 19.71 (3.91) | 18.94 (3.27) | 0.004 |

Values are n (%) unless otherwise stated. *P* values from *t*-test (continuous) or chi-square test (categorical). *SD*, standard deviation; *BMI*, body mass index; *6MWT*, 6-minute walk test; *HIV*, human immunodeficiency virus.

### Nutritional support delivered

Of the 450 recipients, 389 were outpatients and 61 hospitalised. Among the 369 whose basket contents were quantified, the basket provided a median of 3,817 kcal (IQR 2,505– 5,512) during the intensive phase, with a median of 148 g of protein, and was most often received approximately monthly, followed by weekly and fortnightly (Table 2, Supplementary Table 5). The basket was shared among a median of three household members. Delivery patterns were largely site-specific, and receipt among Beninese participants fell from close to 100% early in enrolment to approximately 72% by mid-2024 as programme financing fluctuated.

**Table 2.** Nutritional support delivered to outpatient recipients during the intensive phase.

| Component | Median (IQR) |
| --- | --- |
| Total energy, kcal | 3817 (2505–5512) |
| Protein, g | 148 (82–208) |
| Carbohydrate, g | 505 (256–797) |
| Fat, g | 120 (69–186) |
| Household members sharing | 3 (1–5) |

*IQR*, interquartile range. Cumulative amounts recorded during the intensive phase among 369 outpatient recipients.
Inpatients received prepared meals and are not shown.

### Nutritional and functional outcomes

Unadjusted associations are shown in Supplementary Table 6. Recipients were more likely to achieve ≥5% weight gain at month 2 (45.3% vs 31.7%, p<0.001) and ≥10% weight gain at month 6 (50.4% vs 32.7%, p<0.001). Recipients also gained more weight at month 2 (mean 2.2 vs 1.7 kg, p=0.03) and month 6 (5.3 vs 3.9 kg, p<0.001). After adjustment, receipt of nutritional support was associated with higher odds of ≥5% weight gain at month 2 (aOR 1.57, 95% CI 1.13–2.19) and ≥10% weight gain at month 6 (aOR 1.92, 95% CI 1.35–2.74) (Table 3). Recipients also had greater improvement in 6-minute walk distance at month 6 (adjusted β 40.6 m, 95% CI 26.5 to 54.6).In absolute terms, recipients gained more weight than non-recipients at both month 2 (mean 2.3 vs 1.6 kg, p=0.003) and month 6 (mean 5.3 vs 3.9 kg, p<0.001). Recipients also had a greater increase in their 6MWT at month 6 compared to non-recipients after adjusting for covariates including baseline 6MWT distance.

**Table 3.** Multivariable analysis of nutritional and functional outcomes.

| Outcome | aOR or aβ (95% CI) | P value |
| --- | --- | --- |
| $\geq 5\%$ weight gain, month 2<sup>a</sup> | 1.57 (1.13–2.19) | 0.007 |
| $\geq 10\%$ weight gain, month 6<sup>b</sup> | 1.92 (1.35–2.74) | <0.001 |
| 6MWT change, month 6, m<sup>c</sup> | 40.6 (26.5 to 54.6) | <0.001 |

aOR, adjusted odds ratio; aβ , adjusted regression coefficient; CI, confidence interval; BMI, body mass index; 6MWT, 6-minute walk test. <sup>a</sup> aOR derived from a logistic regression model which controlled for age, sex, multidimensional poverty, ever smoking, hazardous alcohol use, HIV status, and baseline BMI. <sup>b</sup> aOR derived from a logistic regression model which controlled for age, sex, multidimensional poverty, diabetes, and baseline BMI. <sup>c</sup> aβ derived from a linear regression model which controlled for age, sex, HIV status, baseline BMI, and baseline 6MWT distance; baseline distance was included so that the estimate represents the between-group difference in change from baseline to month 6.

### Treatment outcomes

Recipients had higher odds of adherence (aOR 3.43, 95% CI 1.81–6.51). Sputum smear conversion (aOR 0.87, 95% CI 0.50–1.52) and treatment success (aOR 1.37, 95% CI 0.87–2.18) did not differ significantly (Table 4). Mortality was lower among recipients (1.1% vs 3.8%; unadjusted OR 0.29, 95% CI 0.10–0.82), and the association persisted after adjustment (aOR 0.32, 95% CI 0.11–0.93; Supplementary Table 1).

**Table 4.** Multivariable analysis of treatment outcomes.

| Outcome | aOR (95% CI) | P value |
| --- | --- | --- |
| Adherence<sup>a</sup> | 3.43 (1.81–6.51) | <0.001 |
| Sputum smear conversion<sup>b</sup> | 0.87 (0.50–1.52) | 0.623 |
| Treatment success<sup>c</sup> | 1.37 (0.87–2.18) | 0.178 |
| Mortality<sup>d</sup> | 0.32 (0.11–0.93) | 0.036 |

*aOR, adjusted odds ratio; CI, confidence interval; BMI, body mass index. <sup>a</sup> aOR derived from a logistic regression model which controlled for age, sex, ever smoking, and HIV status. <sup>b</sup> aOR derived from a logistic regression model which controlled for age, sex, baseline BMI, and cavitory disease. <sup>c</sup> aOR derived from a logistic regression model which controlled for age, sex, ever smoking, baseline BMI, cavitory disease, and pleural effusion. <sup>d</sup> aOR derived from a logistic regression model which controlled for age and sex. Sputum conversion, treatment success, and mortality analyses involved few events and are exploratory.*

### Subgroup analysis

Weight gain by group is shown in Supplementary Figure 2. Adherence was higher in Benin recipients than non-recipients (96.7% vs 79.6%, p<0.001). With Benin recipients as reference, Benin non-recipients had lower odds of adherence (aOR 0.14, 95% CI 0.06– 0.33) and higher mortality (aOR 6.05, 95% CI 1.38–26.51). For weight gain they did not differ from recipients (≥5% at month 2, aOR 0.93, 95% CI 0.47–1.84), whereas Togo participants were significantly worse (aOR 0.59, 95% CI 0.42–0.84). For 6MWT change, Benin non-recipients did not differ from Benin recipients (aβ 4.6 m, 95% CI −22.4 to 31.6) while Togo participants improved less (aβ −50.7 m, 95% CI −65.5 to −35.8) (Supplementary Table 4).

## DISCUSSION

INSTITUT is the largest programmatic assessment of nutritional support within routine TB care in West Africa, and it addresses a longstanding evidence gap. Programmatic in-kind nutritional support was associated with greater weight gain, better functional recovery, higher adherence, and lower mortality during TB treatment. The absence of a detectable effect on sputum conversion and treatment success most likely reflects limited power and the fact that these outcomes are driven mainly by antimicrobial therapy.

Nutritional support probably acts through two mechanisms. The first is physiological, through effects on immunometabolism, pharmacokinetics, and toxicity risk. The second is behavioural: as a form of social protection, it defrays the cost of care and supports engagement with treatment. Our findings support improvement in nutritional recovery consistent with previous analyses of food baskets and ready to use therapeutic foods including in West Africa [5,11–13], but the extra calories reaching outpatients, after accounting for household sharing, were modest and are unlikely on their own to have produced the weight gain we observed. The weight gain is more plausibly attributable to improved adherence, and possibly to reductions in gastrointestinal adverse effects [14], than to the caloric content of the basket alone. Nutritional support thus acts as an enabler to treatment [12,15–17].

The improvement in 6MWT distance among recipients is also striking, and consistent with qualitative research in which recipients credited nutritional support with improved functional strength during treatment [18]. Underweight PWTB are at increased risk of post-tuberculosis lung disease (PTLD) [19]. Atrophy of intercostal muscles in PWTB who experience cachexia may play a role in this process [20]. Nutritional reconstitution may then mitigate PTLD. Clinical trials are needed to validate this connection and to identify optimal basket composition for lung health. No minimum clinically important difference has been established for TB, but the magnitude we observed exceeds the thresholds of roughly 25 to 30 metres considered clinically relevant in other chronic lung diseases.

Mortality was lower among recipients, and the association persisted after adjustment. Although deaths were few, the direction and magnitude are consistent with the established relationship between undernutrition and TB mortality. To situate this, we pooled INSTITUT with the macronutrient trials reporting mortality in the Grobler Cochrane review (four trials), an additional randomised trial, and a recent cluster-randomised study [5,21–26]. Across all studies the pooled risk ratio favoured nutritional support but included the null (RR 0.56, 95% CI 0.27 to 1.16); excluding the single stepped-wedge study whose mortality ran counter to its own favourable treatment outcomes, the estimate was significant and homogeneous (RR 0.35, 95% CI 0.18 to 0.71; I²=0%) (Figure 2). INSTITUT is the only individual study in this pool whose confidence interval excludes the null, which reflects its size relative to the earlier trials. This synthesis, which combines studies differing in intervention type and outcomes assessed, is intended to place our result in context rather than as a systematic review, and should be regarded as hypothesis-generating.

**Figure 2.**
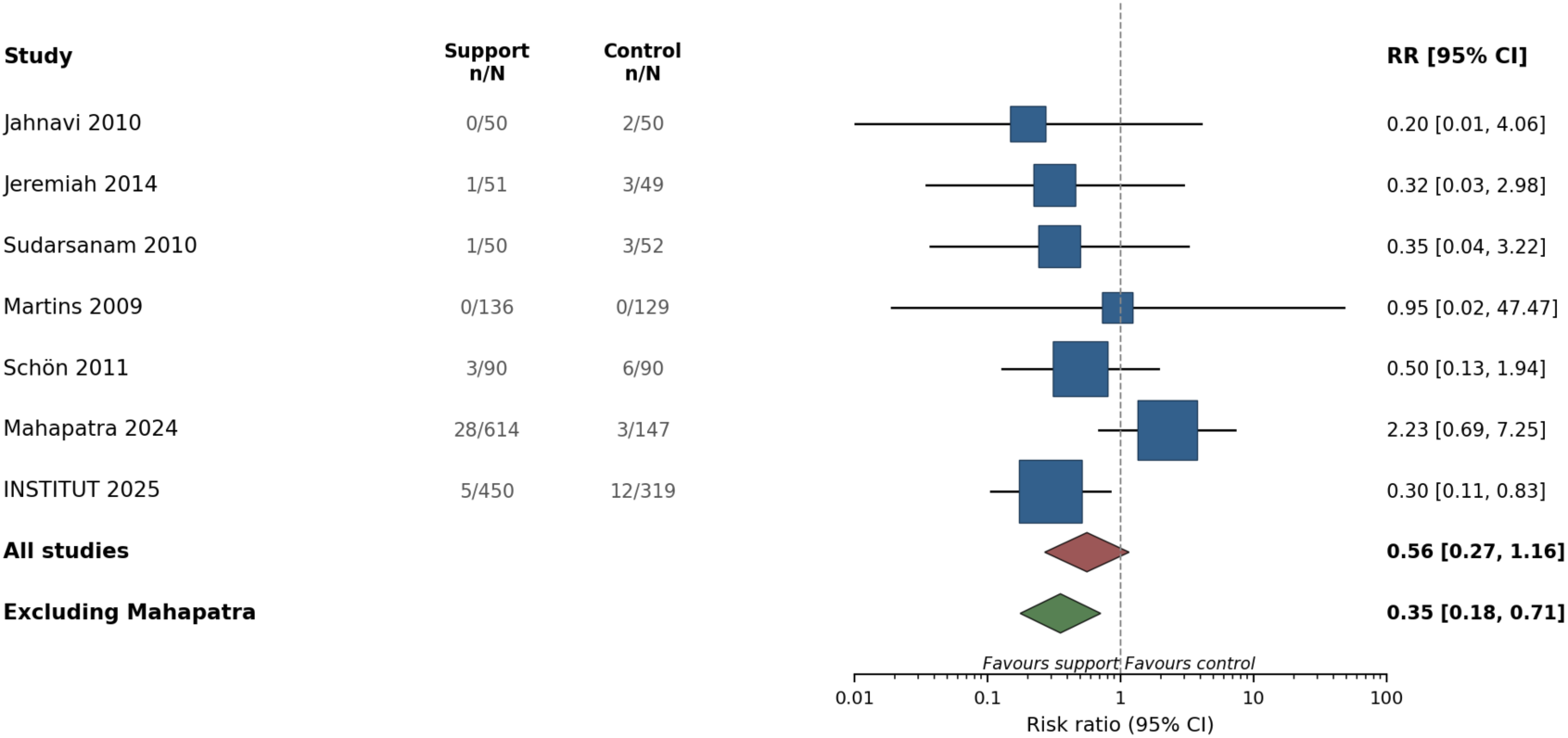
Mortality during tuberculosis treatment: INSTITUT pooled with macronutrient trials reporting mortality. Risk ratios for death during TB treatment. INSTITUT contributes the crude risk ratio (0.30, 95% CI 0.11 to 0.83) so that the effect measure is consistent across studies; this differs slightly from the age- and sex-adjusted odds ratio reported in the main results (0.32, 95% CI 0.11 to 0.93). Arm-level death counts for Jahnavi 2010, Jeremiah 2014, Sudarsanam 2010, and Martins 2009 are from the Cochrane review (Grobler 2016); Schön 2011 from the primary report; Mahapatra 2024 from the source study. Estimates were pooled by random-effects meta-analysis (DerSimonian-Laird), with marker size proportional to study weight. Between-study heterogeneity was low (all studies I²=23%; excluding Mahapatra I²=0%).

The subgroup analysis provides an internal check. Benin non-recipients, who shared Benin’s health system and sites but received no food support, had lower adherence and higher mortality than recipients, and were directionally worse for weight gain. Because they differed only in whether supply was available, this is difficult to explain by country-level factors. The exception is the 6-minute walk test, where Benin non-recipients improved as much as recipients while Togo participants improved much less, so the functional advantage over Togo may partly reflect country-level differences rather than nutritional support alone. Benin non-recipients numbered only 49, so these subgroup estimates are imprecise.

Our study has limitations. The non-randomised design leaves residual confounding possible, particularly from unmeasured differences between Benin and Togo; the subgroup analysis mitigates but does not eliminate this concern. Support was heterogeneous in quantity and schedule, organised largely by site, and shared within households; receipt declined as programme financing fluctuated, which is why some Beninese participants received none. This heterogeneity, with enrolment below the design target, left us underpowered to examine dose-response and to detect differences in sputum conversion and treatment success. Outcome data were incomplete for some participants, with higher missingness among Togo participants and non-recipients.

Seventeen enrolled participants transferred out before outcomes could be ascertained; they were comparable to those analysed except for lower baseline walk distance (315 vs 382 m, p=0.03) and were predominantly from Togo, so their exclusion would bias the walk-distance comparison against nutritional support. Receipt was ascertained from programme records and participant report, so misclassification is possible, although a sensitivity analysis restricting recipients to those with quantified basket contents gave similar estimates.

Despite these limitations, the convergent findings across nutritional, functional, and behavioural outcomes, together with the subgroup analysis, support the conclusion that a programmatic in-kind food basket provided alongside standard TB treatment is operationally feasible and clinically meaningful. While the causal nature of these associations requires further exploration, nutritional support appears to be a person-centred intervention that supports functional and nutritional recovery and may also help TB programmes recoup their investment in diagnosis and treatment by encouraging people with TB to complete therapy.

These findings provide a compelling impetus for adopting similar programmes across the African continent. Addressing the nutritional dimension of TB with the same urgency applied to HIV and diabetes is both a clinical priority and an ethical imperative.

## DATA AVAILABILITY

Data are available upon reasonable request to the corresponding author.

## ACKNOWLEDGEMENTS

We acknowledge the tremendous contributions of our field teams in Benin and Togo for working assiduously and sincerely to collect these multicenter data that inform action for vulnerable individuals in West Africa.

## Supplementary Material

**Supplementary Table 1.**
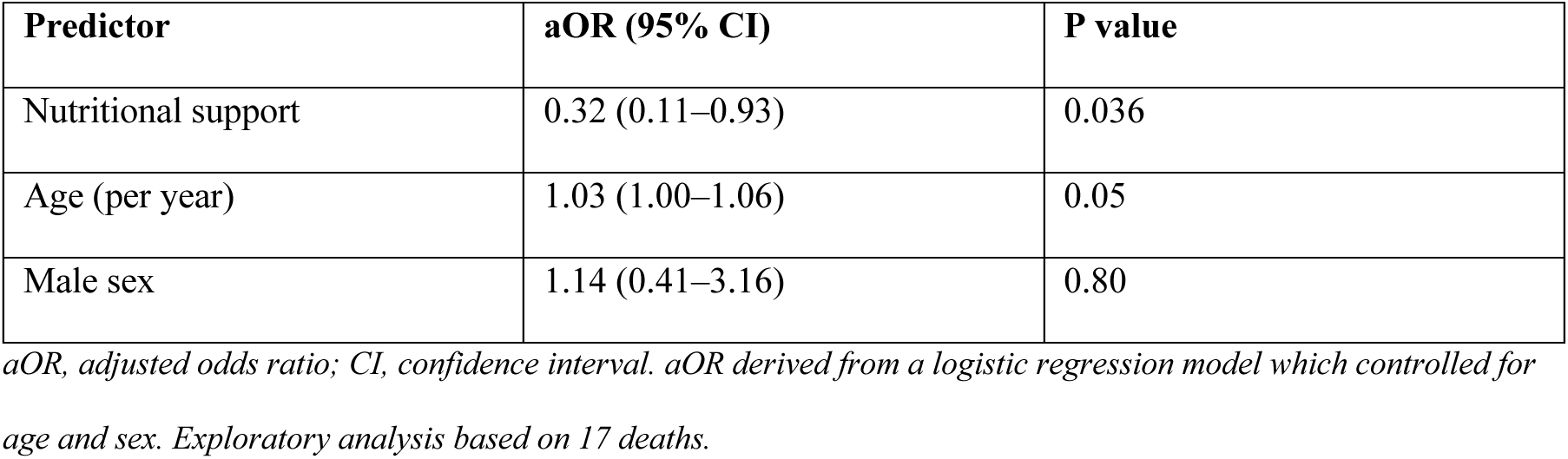
Multivariable analysis of mortality.

**Supplementary Table 2.**
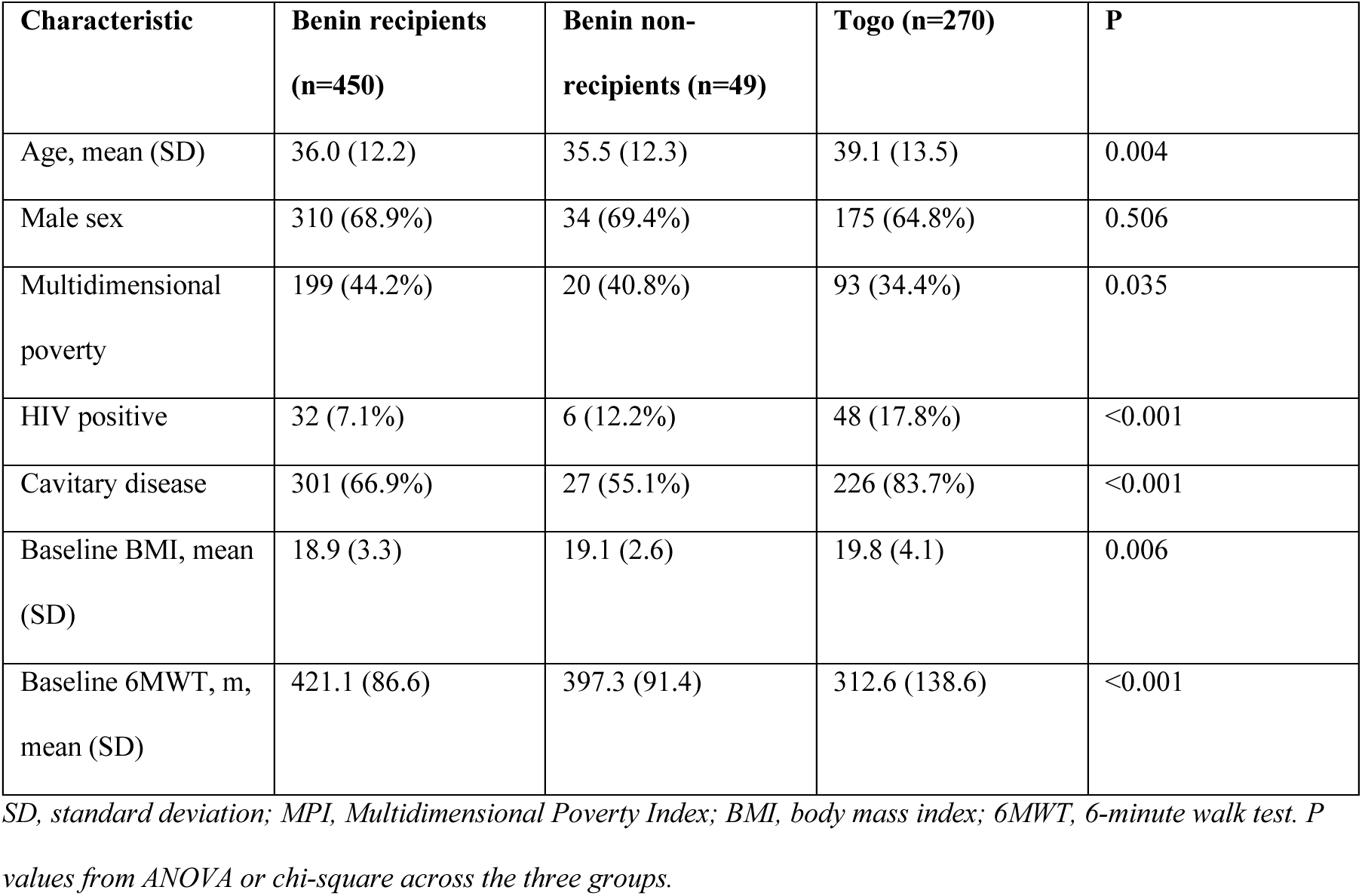
Baseline characteristics by group.

**Supplementary Table 3.** Missing data by outcome and group.

| Outcome | Benin recipients | Benin non-recipients | Togo | Total |
| --- | --- | --- | --- | --- |
| Weight, month 2 | 4 (0.9%) | 9 (18.4%) | 17 (6.3%) | 30 (3.9%) |
| Weight, month 6 | 53 (11.8%) | 11 (22.4%) | 42 (15.6%) | 106 (13.8%) |
| 6MWT, month 6 | 56 (12.4%) | 11 (22.4%) | 49 (18.1%) | 116 (15.1%) |
| Smear conversion, month 2 | 134 (29.8%) | 22 (44.9%) | 77 (28.5%) | 233 (30.3%) |
| Treatment outcome | 0 (0.0%) | 0 (0.0%) | 0 (0.0%) | 0 (0.0%) |

Values are n (%). Adherence and treatment outcome were complete for all analysed participants.

**Supplementary Table 4.**
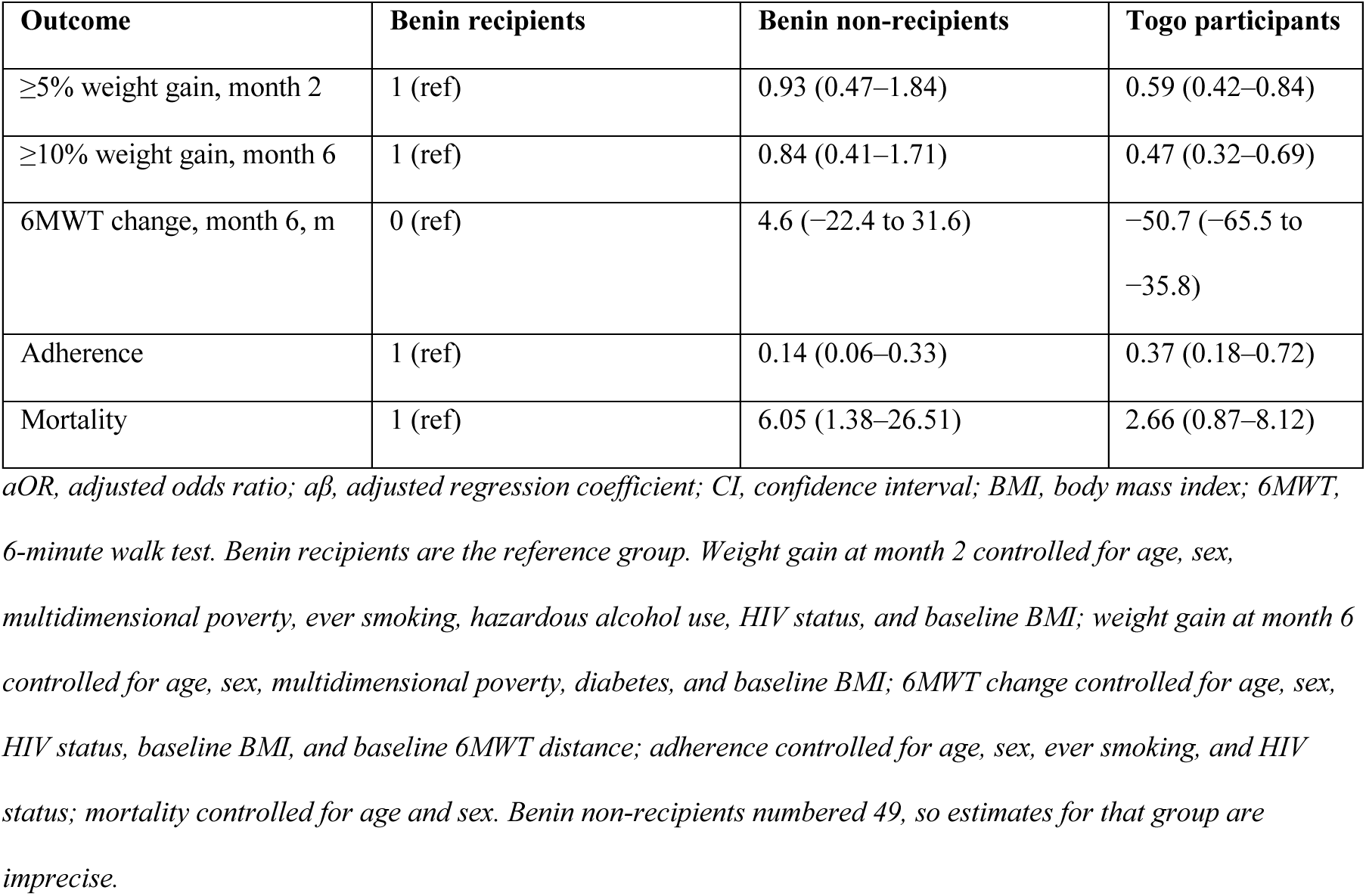
Multivariable analysis of adherence by subgroup.

**Supplementary Table 5.**
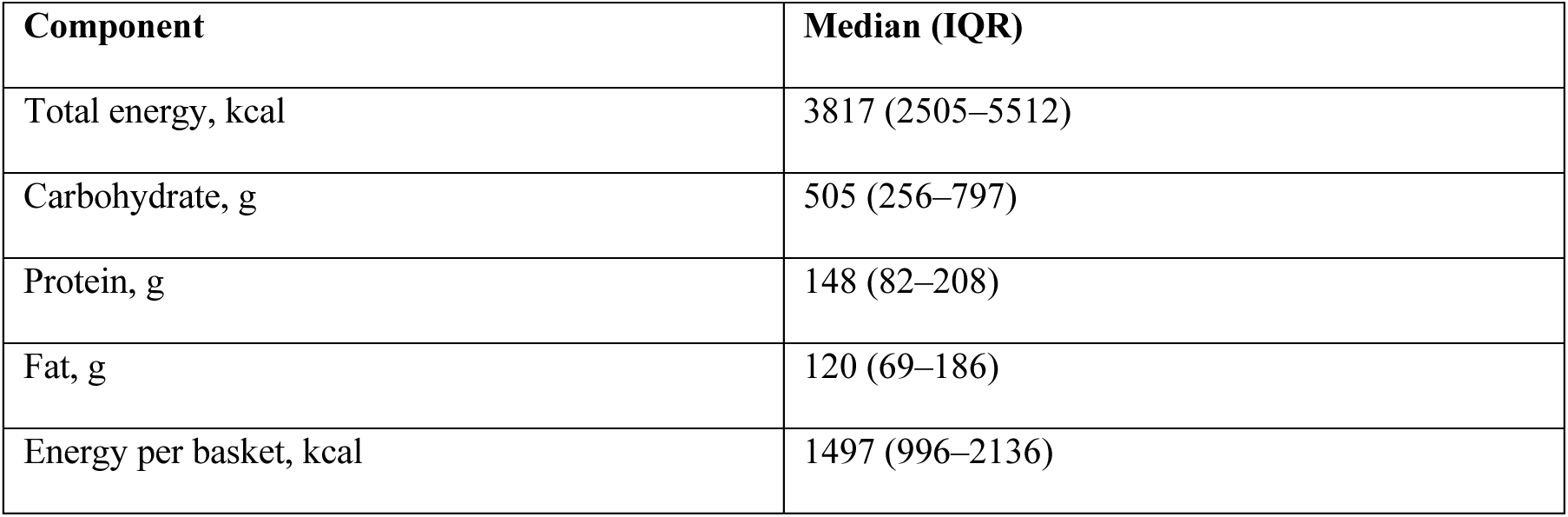
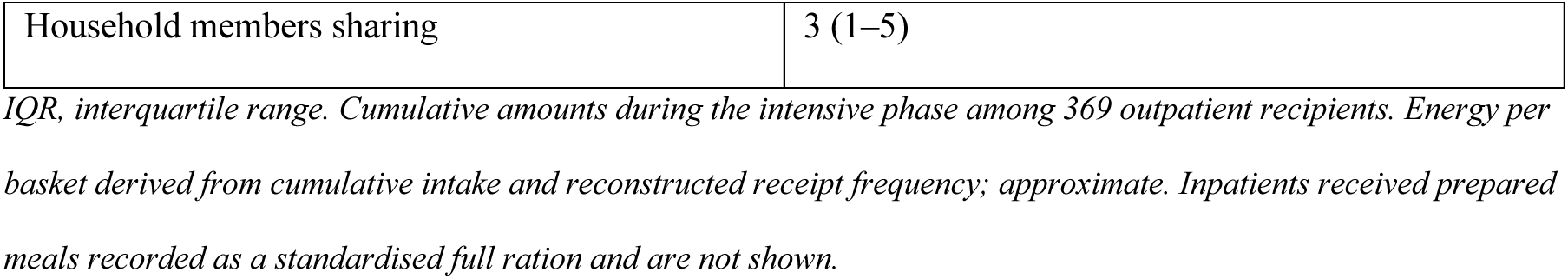
Cumulative nutritional support delivered to outpatient recipients.

| Component | Median (IQR) |
| --- | --- |
| Total energy, kcal | 3817 (2505–5512) |
| Carbohydrate, g | 505 (256–797) |
| Protein, g | 148 (82–208) |
| Fat, g | 120 (69–186) |
| Energy per basket, kcal | 1497 (996–2136) |
| Household members sharing | 3 (1–5) |

*IQR, interquartile range. Cumulative amounts during the intensive phase among 369 outpatient recipients. Energy per basket derived from cumulative intake and reconstructed receipt frequency; approximate. Inpatients received prepared meals recorded as a standardised full ration and are not shown.*

**Supplementary Table 6.** Univariable (unadjusted) associations between receipt of nutritional support and study outcomes.

| Outcome | Unadjusted OR or $\beta$ (95% CI) | P value |
| --- | --- | --- |
| $\geq 5\%$ weight gain, month 2 | 1.78 (1.31–2.42) | <0.001 |
| $\geq 10\%$ weight gain, month 6 | 2.09 (1.51–2.89) | <0.001 |
| 6MWT change, month 6, m | 3.5 (–11.3 to 18.3) | 0.642 |
| Adherence | 3.23 (1.72–6.08) | <0.001 |
| Sputum smear conversion | 0.84 (0.49–1.45) | 0.539 |
| Treatment success | 1.33 (0.85–2.08) | 0.207 |
| Mortality | 0.29 (0.10–0.82) | 0.020 |

*OR, odds ratio (logistic regression); $\beta$ , regression coefficient (linear regression) for 6-minute walk test (6MWT)*
*change; CI, confidence interval. Estimates are unadjusted associations with receipt of nutritional support. Adjusted estimates are shown in Tables 3 and 4. The univariable 6MWT estimate is smaller than the adjusted estimate because Togo participants had substantially lower baseline 6MWT distance; adjustment for baseline distance accounts for this.*

**Supplementary Figure 1.**
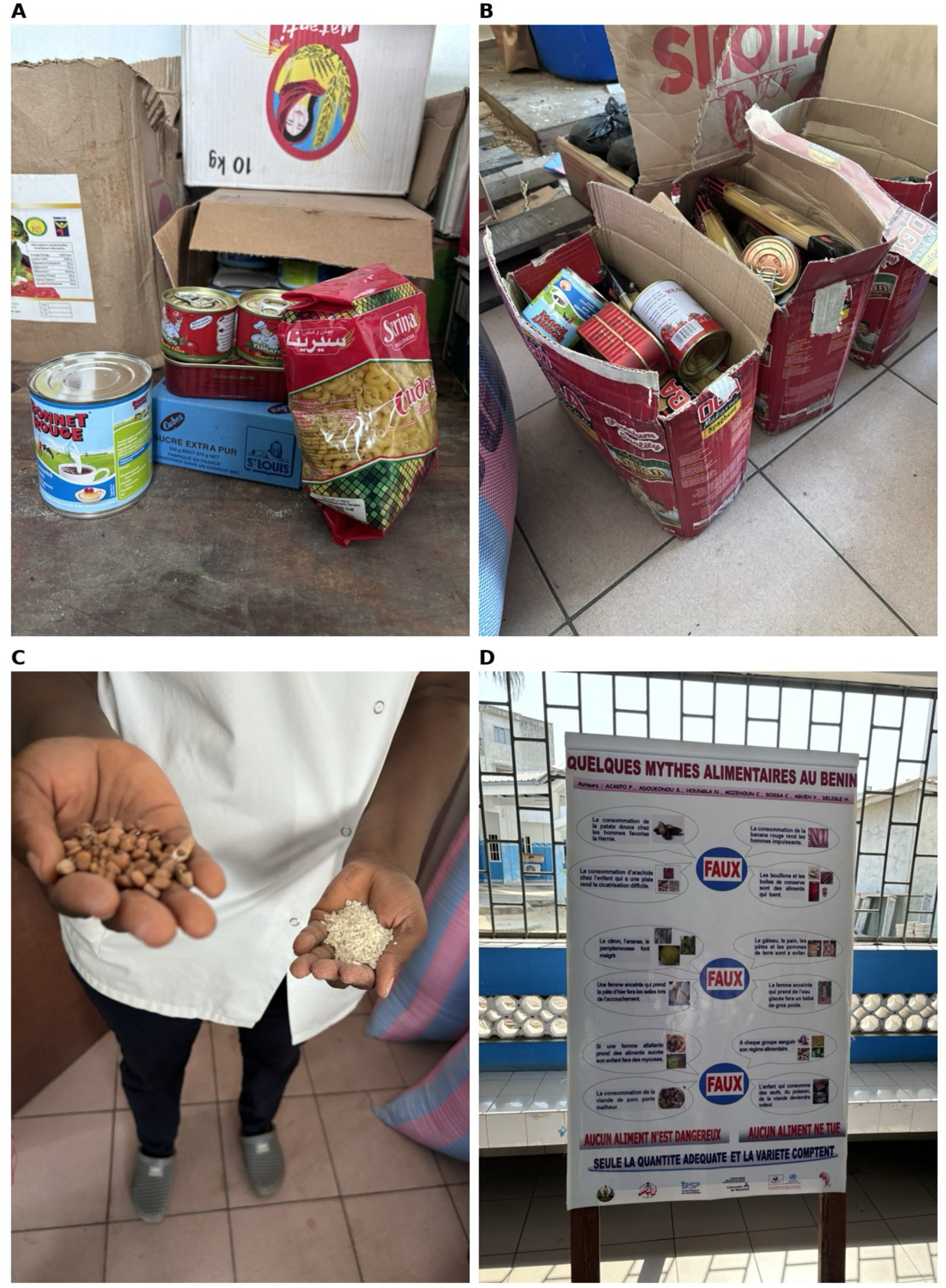
Programmatic nutritional support provided to people with tuberculosis in Benin. (A) Assembled contents of a monthly take-home food basket, including durum-wheat pasta, tinned sardines and corned beef, sweetened condensed milk, sugar, and tomato paste. (B) Baskets prepared for distribution. (C) Rice and dried legumes provided in the basket. (D) Nutritional counselling poster used at follow-up visits, addressing local dietary practices and food misconceptions.

**Supplementary Figure 2.**
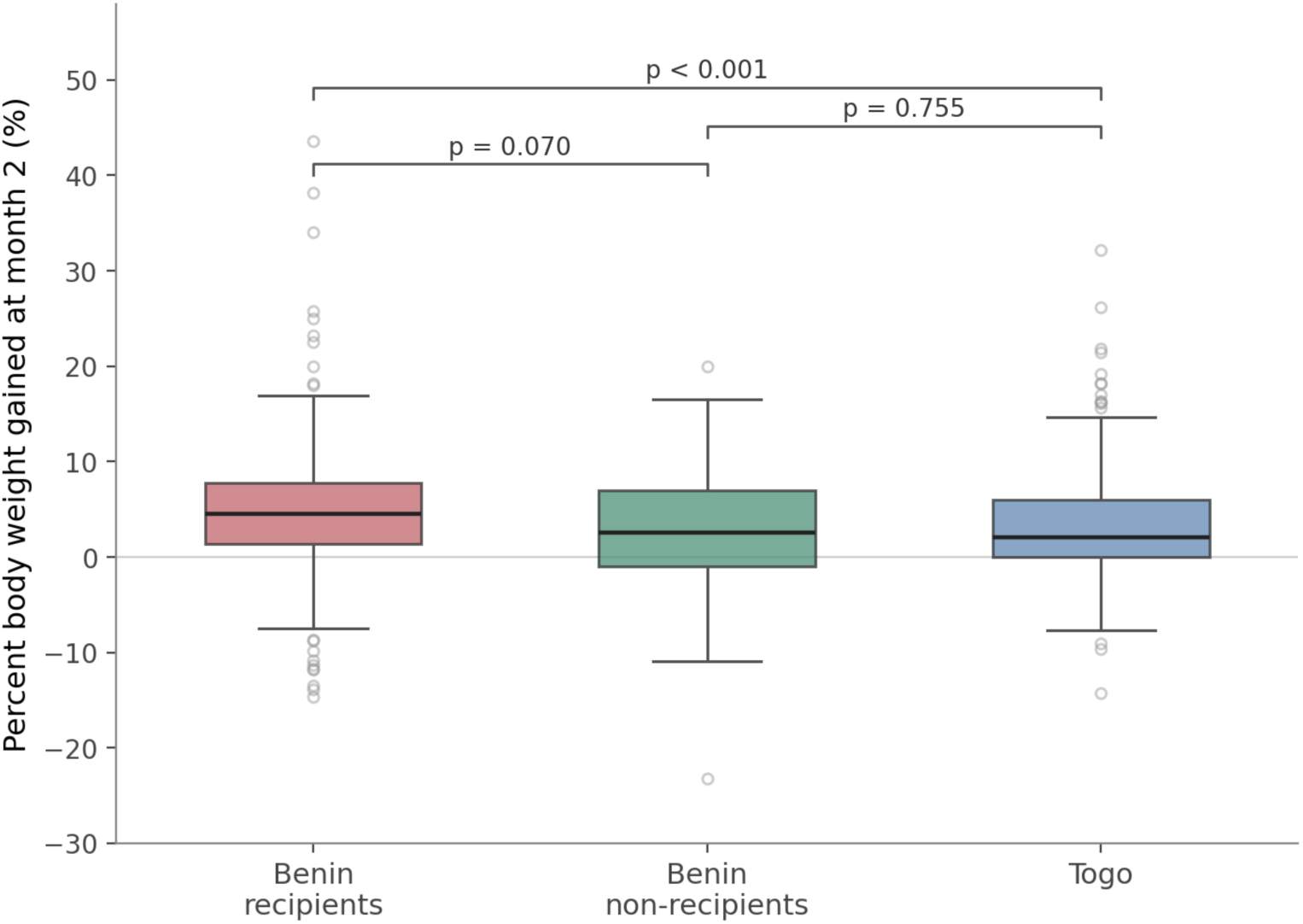
Percent body weight gained at month 2 by group (Benin recipients, Benin non-recipients, Togo).

## REFERENCES

1. World Health Organization. Global tuberculosis report. Geneva: WHO. [confirm edition/year]

2. Sinha P, Davis J, Saag L, Wanke C, Salgame P, Mesick J, Horsburgh CR Jr, Hochberg NS. Undernutrition and tuberculosis: public health implications. J Infect Dis. 2019;219(9):1356–63.

3. Tabackman A, Gupte AN, Thangakunam B, Babu SP, Du X, Paradkar M, et al. [Premorbid nutritional status and TB severity]. Int J Tuberc Lung Dis. 2025. [confirm full citation]

4. Sinha P, White LF, Hochberg NS, Cegielski JP. Avoiding pitfalls in calculating the population attributable fraction of undernutrition for TB. Int J Tuberc Lung Dis. 2022;26(1):80.

5. Grobler L, Nagpal S, Sudarsanam TD, Sinclair D. Nutritional supplements for people being treated for active tuberculosis. Cochrane Database Syst Rev. 2016;(6):CD006086.

6. Dauphinais MR, Koura KG, Narasimhan PB, Mehta S, Finkelstein JL, Heysell SK, Sinha P. Nutritionally acquired immunodeficiency must be addressed with the same urgency as HIV to end tuberculosis. BMC Glob Public Health. 2024;2(1):4.

7. Cintron C, Dauphinais M, Fiogbe AA, Dogo MF, Ate S, Adjonou C, et al. Impact of Nutritional Support for Tuberculosis on Intermediate and Terminal Undernutrition and Treatment Outcomes (INSTITUT) study: a protocol. BMJ Open. 2025;15:e088629.

8. World Bank. Gini index. Washington, DC: World Bank. https://data.worldbank.org/indicator/SI.POV.GINI

9. ATS Committee on Proficiency Standards for Clinical Pulmonary Function Laboratories. ATS statement: guidelines for the six-minute walk test. Am J Respir Crit Care Med. 2002;166(1):111–7.

10. Cintron C, Dauphinais MR, Du X, Tabackman A, Lenart A, Laliberte A, Dirksen J, Sinha P. Enriching tuberculosis research by measuring poverty better: a perspective. BMC Glob Public Health. 2025;3(1):17.

11. Kumar R, Sinha P, Krishnan A, Singh M, Singh A, Guleria R, Singh UB. Energy dense nutritional supplements improve weight gain among malnourished adults with drug-sensitive pulmonary tuberculosis: an open-label randomized controlled trial in Faridabad, India. Int J Infect Dis. 2026;163:108286.

12. Sinha P, Bhargava M, Carwile ME, Dauphinais MR, Tisile P, Cintron C, et al. A roadmap for integrating nutritional assessment, counselling, and support into the care of people with tuberculosis. Lancet Glob Health. 2025;13:e967–73.

13. Bhargava A, Bhargava M, Meher A, Teja GS, Velayutham B, Watson B, et al. Nutritional support for adult patients with microbiologically confirmed pulmonary tuberculosis: outcomes in a programmatic cohort nested within the RATIONS trial in Jharkhand, India. Lancet Glob Health. 2023;11(9):e1402–11.

14. Burman WJ, Martins MF, Flynn D, Johnston J, Sinha P, Horsburgh CR. A scoping review of interventions to prevent and treat adverse events during treatment of rifampin-susceptible tuberculosis. PLoS One. 2025;20(12):e0339354.

15. Sinha P, Carwile M, Bhargava A, Cintron C, Acuna-Villaorduna C, Lakshminarayan S, et al. How much do Indians pay for tuberculosis treatment? A cost analysis. Public Health Action. 2020;10(3):110–7.

16. Wagnew F, Gray D, Tsheten T, Kelly M, Clements ACA, Alene KA. Effectiveness of nutritional support to improve treatment adherence in patients with tuberculosis: a systematic review. Nutr Rev. 2024;82(9):1216–25.

17. Benzekri NA, Sambou JF, Tamba IT, Diatta JP, Sall I, Cisse O, et al. Nutrition support for HIV-TB co-infected adults in Senegal, West Africa: a randomized pilot implementation study. PLoS One. 2019;14(7):e0219118.

18. Carwile M, Jain K, Dauphinais M, Cintron C, Janarthanan, Locks LM, Maloomian K, Prakash Babu S, Rajkumari N, R M, Narasimhan PB. Tuberculosis-learning about experience with nutritional supplementation (TB LENS): perspectives on nutritional supplementation for persons with TB and their household contacts. Glob Public Health. 2025;20(1):2576758.

19. Sinha P, Karoly M, Gaikwad S, Hanna LE, Gupte N, Kulkarni V, Pradhan N, Suryavanshi N, Dauphinais MR, Zhang V, Selvaraju S. The impact of undernutrition on post-tuberculosis lung disease: magnitude and mechanisms. Preprint. Available at SSRN: https://papers.ssrn.com/sol3/papers.cfm?abstract_id=6246933

20. Arora NS, Rochester DF. Respiratory muscle strength and maximal voluntary ventilation in undernourished patients. Am Rev Respir Dis. 1982;126(1):5–8.

21. Sudarsanam TD, John J, Kang G, Mahendri V, Gerrior J, Franciosa M, et al. Pilot randomized trial of nutritional supplementation in patients with tuberculosis and HIV-tuberculosis coinfection receiving directly observed short-course chemotherapy for tuberculosis. Trop Med Int Health. 2011;16(6):699–706.

22. Jahnavi G, Sudha CH. Randomised controlled trial of food supplements in patients with newly diagnosed tuberculosis and wasting. Singapore Med J. 2010;51(12):957–62.

23. Jeremiah K, Denti P, Chigutsa E, Faurholt-Jepsen D, PrayGod G, Range N, et al. Nutritional supplementation increases rifampin exposure among tuberculosis patients coinfected with HIV. Antimicrob Agents Chemother. 2014;58(6):3468–74.

24. Martins N, Morris P, Kelly PM. Food incentives to improve completion of tuberculosis treatment: randomised controlled trial in Dili, Timor-Leste. BMJ. 2009;339:b4248.

25. Schön T, Idh J, Westman A, Elias D, Abate E, Diro E, et al. Effects of a food supplement rich in arginine in patients with smear positive pulmonary tuberculosis: a randomised trial. Tuberculosis (Edinb). 2011;91(5):370–7.

26. Mahapatra A, Thiruvengadam K, Nair D, Padmapriyadarsini C, Thomas B, Pati S, et al. Effectiveness of food supplement on treatment outcomes and quality of life in pulmonary tuberculosis: phased implementation approach. PLoS One. 2024;19:e0305855.

